# Duration of viral shedding of the Omicron variant in asymptomatic and mild COVID-19 cases from Shanghai, China

**DOI:** 10.1101/2022.12.08.22283272

**Authors:** Weijie Sun, Naibin Yang, Yang Mao, Danying Yan, Qifa Song, Guoqing Qian

**Author notes:** **Corresponding authors;** Guoqing Qian, Qifa Song.

## Abstract

**Background:** The Omicron variant of severe acute respiratory syndrome coronavirus 2 (SARS-CoV-2), designated as a variant of concern by the World Health Organization, spreads globally and was confirmed as the cause of the Omicron wave of the coronavirus disease 2019 (COVID-19) pandemic in Shanghai, China. The viral shedding duration of Omicron variants needs to be determined.

**Methods:** We retrospectively analyzed 382 patients admitted to a shelter hospital for COVID-19. Of the patients, 8 patients were referred to a designated hospital, 100 were infected asymptomatic patients, and 274 patients had mild COVID-19.

**Results:** The vaccination rates (including fully and boosted) in the asymptomatic and mild COVID-19 patients were 92.00% and 94.16%, respectively. Majority of the studied population showed a first reverse transcription-polymerase chain reaction cycle threshold (Ct) value of 20. For 2565 nasopharyngeal swabs from close or sub-close contacts, the Ct value gradually increased to 35 for 8 days, and the median duration of viral shedding time was 10 days after the first positive detection of the SARS-CoV-2 nuclei acid.

**Conclusions:** Quantitative viral RNA load assays in COVID-19 (BA.2.2.1) close or sub-closed contacts could be used to prevent transmissions and control precautions.

## Introduction

Omicron variants of severe acute respiratory syndrome coronavirus 2 (SARS-CoV-2) have spread rapidly worldwide, causing outbreaks with 63,007 confirmed cases and 595 deaths in Shanghai, China from March to June 2022 [1]. Studies have indicated lower virulence and lethality of Omicron variants than of Delta variants. Along with pneumonia, runny nose, headache, and fatigue are the most common symptoms of Omicron [2; 3]. However, the sequelae and long-term effects of this variant remain unclear. Especially in China, although inactivated vaccines have been widely used, Omicron variants may still be a huge public health concern in a country with the largest population and highly aging rate [4].

The spread of Omicron variant in Shanghai communities has been well controlled. Even after this wave in Shanghai, many challenges exist in terms of prevention and treatment of Omicron variants. Measures such as isolation have proved effective in suppressing the spread of the Omicron variants in China [5]. However, the economy, society, and medical staff have paid a huge price during lockdown conditions. We need to constantly accumulate and update our knowledge about coronavirus disease 2019 (COVID-19), thereby improving our isolation measures and implementing the most suitable treatment method to control the spread of the Omicron variant. Moreover, the Omicron variants might dynamically be mutated during infection replications. A systemic and dynamic study of the virology of Omicron variants, particularly in China, is lacking[6]. Therefore, to provide a real-world data analysis for patient isolation, we here analyzed the duration of viral shedding of the Omicron variants in infected asymptomatic and mild COVID-19 cases from Shanghai in a shelter hospital.

## Materials and Methods

This retrospective study was conducted at the Cixi Dapengshan Shelter Hospital (managed by Ningbo First Hospital, University of Ningbo) and was approved by the Ethics Commission of Ningbo First Hospital (approval No. 2022RS065). Written informed consent was waived because of the study’s retrospective nature. Data of 382 consecutive asymptomatic or mild COVID-19 individuals were investigated, and no patient was classified as severe or critical case at hospital admission. These patients were from Shanghai City and were recruited from Cixi Dapengshan Shelter Hospital between April 4 and April 28, 2022.

Methods for construction and determination of the whole genome library were as followed. Total RNA was extracted from 200 μL samples using a viral nucleic acid extraction Kit (Qiagen RNeasy Mini Kit/74104); multiple amplification products were obtained by ultra-sensitive SARS-CoV-2 whole genome capture kit (Microfuture /V-090418); AMPure XP beads/A63880 were used to purify the amplification products; Qubit dsDNA HS ASSAY Kit/1646715 was used for quantification; the auxiliary library construction kit (Hangzhou Baiyi Technology Co., LTD. /BK-AUX024) was used for library construction; the connector sequencing kit (Oxford Nanopore Technologies/SQK-LSK110) was used for connection and on-machine preparation, and the sequencing chip (Oxford Nanopore Technologies/ FLO-MIN106) was used for high-throughput sequencing.CLC was used to concatenate the data and generate consensus sequences. The reference genome was Wuhan-1 (Genbank Accession No.NC045512.2). The sequences were typed using Pangolin typing (Pangolin version: v2.3.1).

The first day of testing positive for the Omicron variant was defined as the diagnosis date from close or sub-close contacts. The sample for reverse transcription-polymerase chain reaction (RT-PCR) was collected using nasopharyngeal swabs before admission and during hospitalization in the shelter hospital. These nasopharyngeal samples were collected from 382 patients confirmed with COVID-19. Real-time RT-PCR was performed, and the presence of the SARS-CoV-2 nucleocapsid gene (N and ORF genes) was confirmed [7].This was the first RT-PCR was, using the following two types of kits, according to the manufacturer’s instructions: Shengxiang (Shengxiang Biotechnology, Hunan, China) and Mingde (Wuhan Easy Diagnosis Biomedicine, Wuhan, China). As operation principle, the multiplex PCR-fluorescent probe technology combined with one-step RT-PCR technology was used in the Mingde test kit while paramagnetic particle sorting method without nucleic acid extraction combined with one-step RT-PCR technology was used in the Shengxiang test kit. No matter for Mingde or Shengxiang kit, the specific conserved sequence of SARS-CoV-2 virus ORF 1ab and encoding nucleocapsid protein N gene was used as the target region, and the two-target gene detection primer probe was designed, and the RT-PCR reaction system was used for detection. After the reaction, the threshold cycle number (Ct value) of each channel was analyzed. At the same time, the human housekeeping gene RNaseP was designed as an internal standard to effectively prevent the occurrence of false positives and false negatives to ensure the accuracy of the test results.

According to the national guidelines on COVID-19 diagnosis and treatment in China (Version No.9) [8], cycle threshold (Ct) value of ≥35 must be obtained following two consecutive negative tests (more than 24h) or obtained Ct >40 (negative result, twice in >24h) cycles before being discharged from the shelter hospital and monitored at home [8]. A lower Ct value indicates a higher viral load in the samples and vice versa. Generally, a Ct value of 33 nearly equals 500 viral copies, and 3.3 cycles equal a 10-fold change in viral copies [9].

Data analysis and visualization were performed by R version 4.1.1. The Mann-Whitney t-test by SPSS (25.0) was used, and statistical significance was set at p<0.05.

## Results

### The results of whole genome sequencing of nasopharyngeal samples

The Nanopore third-generation genome was sequenced to these nasopharyngeal samples, and the genome coverage was 99.66%. A total of 74 base site mutations were detected, involving 53 amino acid variations. The results showed the samples belonged to the BA.2.2.1 evolutionary branch (GenBank accession number SUB11780514 NBCDC_20220315_HuYB ON955521).

### Demographic and clinical features of individuals infected by the Omicron variant

All 382 individuals infected by the Omicron variant were from the designated isolation hotel, as confirmed through a positive RT-PCR result and in accordance with the guidelines of the Center for Disease Control and Prevention (CDC). Overall, 165 patients (43.19%) were men, and the patient median age was 32 (26-38) years. Except 7 children and 1 pregnant patient, 374 patients underwent chest computed tomography (CT) after admission. Overall, 100 patients were asymptomatic and 274 were diagnosed with mild COVID-19 (Figure 1). Eight infections were referred to as mild COVID-19 with the emergence of new symptoms. Five patients were diagnosed with moderate COVID-19 on the basis of results of chest CT scans, including 2 asymptomatic patients and 3 patients with mild infections. Moderate COVID-19 was defined as the onset of fever and/or respiratory symptoms and radiographic evidence of pneumonia [8].Among the 382 patients, 1 patient was diagnosed with pulmonary tuberculosis, 1 was diagnosed with acute coronary syndrome due to chest pain, and 1 was diagnosed with the severe COVID-19 risk factor due to systemic lupus erythematosus and the long-term use of corticosteroids. All these 3 patients and the aforementioned 5 individuals with moderate pneumonia were transferred to the designated hospital after diagnosis was established and a risk factor evaluation was completed (Figure 1).

**Figure 1.**
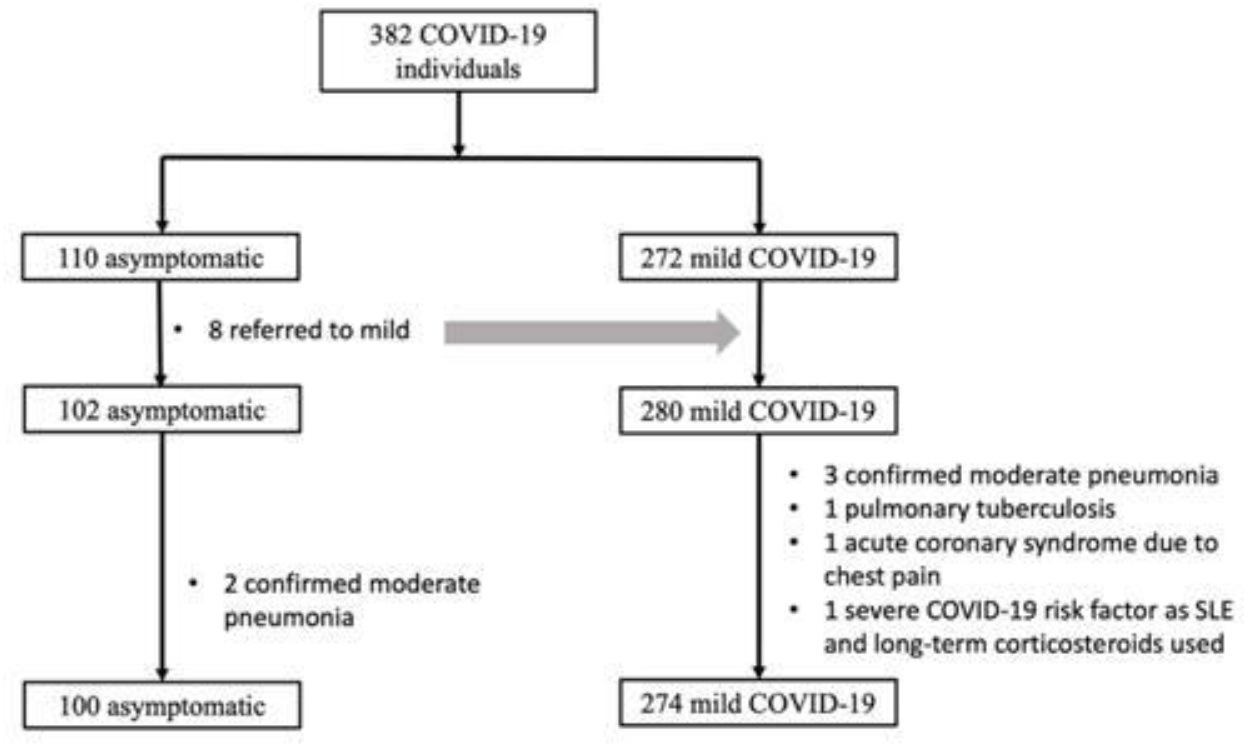
Flowchart shows the enrolment of patients with COVID-19 for this study. Moderate COVID-19 was defined as the onset of fever and/or respiratory symptoms and radiographic evidence of pneumonia. No patients were diagnosed with severe or critical COVID-19 during hospitalization in the shelter hospital.

According to Table 1, 92.00% patients in the asymptomatic group were fully vaccinated or had received booster vaccinations. On the other hand, 41.24% and 51.92% patients in the mild COVID-19 group were fully vaccinated or had received booster vaccinations, respectively. The median duration of viral shedding for the asymptomatic and mild COVID-19 cases was 10 days, and no difference was noted between the two clinical types.

**Table 1.** Comparison of demographic, vaccination, and viral shedding time in patients with asymptomatic and mild COVID-19.

| Variable | Asymptomatic | mild COVID-19 | P-value |
| --- | --- | --- | --- |
| Age (years, median [Q1-Q3]) | 34[29-45] | 31[26-37] | **0.021** |
| Age groups [n (%)] |  |  | 0.268 |
| <18 | 3 (3.00) | 6 (2.19) |  |
| 18-39 | 80 (80.00) | 216 (78.83) |  |
| 40-59 | 14 (14.00) | 42 (15.33) |  |
| ≥60 | 3 (3.00) | 10 (3.65) |  |
| Gender [male, n (%)] | 48(48.00) | 112 (40.88) | 0.239 |
| **Vaccination status [n (%)]** |  |  | 0.100 |
| Not vaccinated | 7 (7.00) | 7 (2.55) |  |
| Partially vaccinated | 1 (1.00) | 9 (3.28) |  |
| Fully vaccinated | 49 (49.00) | 113 (41.24) |  |
| Booster vaccinated | 43 (43.00) | 145 (52.92) |  |
| Discharge [n (%)] |  |  | 0.937 |
| Ct value ≥35 | 53 (53.00) | 145 (52.92) |  |
| Ct value >40 (negative) | 47 (47.00) | 129 (47.08) |  |
| Viral shedding days | 10[9-11] | 10[9-12] | 0.345 |

Notes: The Mann-Whitney t-test was used to compare difference between asymptomatic patients group and mild COVID-19 patients. Statistical significance was set at $p < 0.05$ . These continuous variables were not in accordance with normal distribution and expressed as median [quartile1-quartile 3]. Categorical variables were presented as n (%). Abbreviation: COVID-19, coronavirus disease 2019; Ct, cycle threshold.

### Ct value distribution of first positive RT-PCR

All 382 patients were screened through RT-PCR in the designated isolation hotel or hospital. According to Table 2, two types of kits, Mingde and Shengxiang, were used by the CDC laboratory to detect and confirm SARS-CoV-2 expression. Figure 2 shows the distribution of the Ct value of primary RT-PCR results. The Ct value of approximately 20 patients accounted for the main proportion, with a high copy number of virus in these patients.

**Table 2.** The average CT values of patients at the time of first positive RT-PCR detection.

| Kit Brands | Asymptomatic | mild COVID-19 | P-value |
| --- | --- | --- | --- |
| Mingde (N) | 24.68±5.46 | 25.07±6.59 | 0.762 |
| Mingde (ORF1ab) | 24.17±5.63 | 25.87±6.77 | 0.192 |
| Shengxiang (N) | 23.06±5.68 | 22.98±5.44 | 0.933 |
| Shengxiang (ORF1ab) | 24.38±4.99 | 24.42±5.19 | 0.957 |

Note: The Mann-Whitney t-test was used to compare difference between asymptomatic patients group and mild COVID-19 patients. Statistical significance was set at $p < 0.05$ . Two types of kits, Mingde and Shengxiang, were used to detect and confirm the expression of SARS-CoV-2 by CDC laboratory. All these continuous variables followed in accordance with normal distribution and expressed as Mean ± SD.

**Figure 2.**
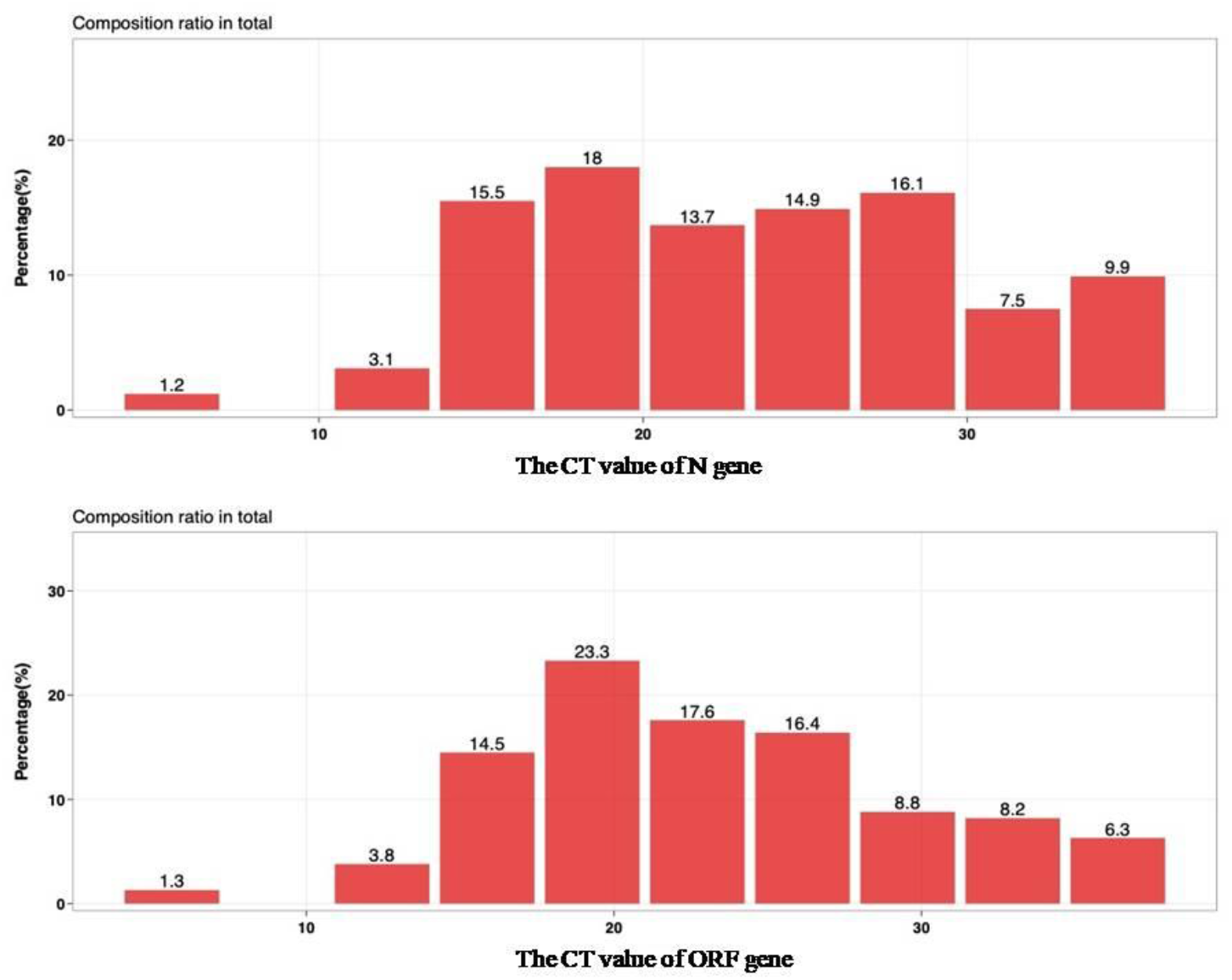
The distribution of the first Ct value for the diagnosis of COVID-19 before admission (Mingde kit: N gene and ORFgene).

### Kinetic viral shedding in Omicron-infected cases with asymptomatic and mild COVID-19

A total of 2565 nasopharyngealswabs were collected from all recruited individuals from close exposure or sub-close exposure to discharge from the hospital. As shown in Figure 3 and 4, the Ct value decreased after admission for 1-2 days and then gradually increased to 35 by approximately day 8. The median duration of viral shedding was 10 days after the first positive detection of SARS-CoV-2. Furthermore, the viral loads soon decreased to the detection limit after day 8. Interestingly, on day 1 of hospitalization, the Ct value in the age group of >50 years was significantly lower than that in the younger age groups (P<0.05). No significant difference in the viral loads was observed across sex (p=0.239), asymptomatic or mild COVID-19 status(p=0.345), and vaccination status (p=0.1).

**Figure 3.**
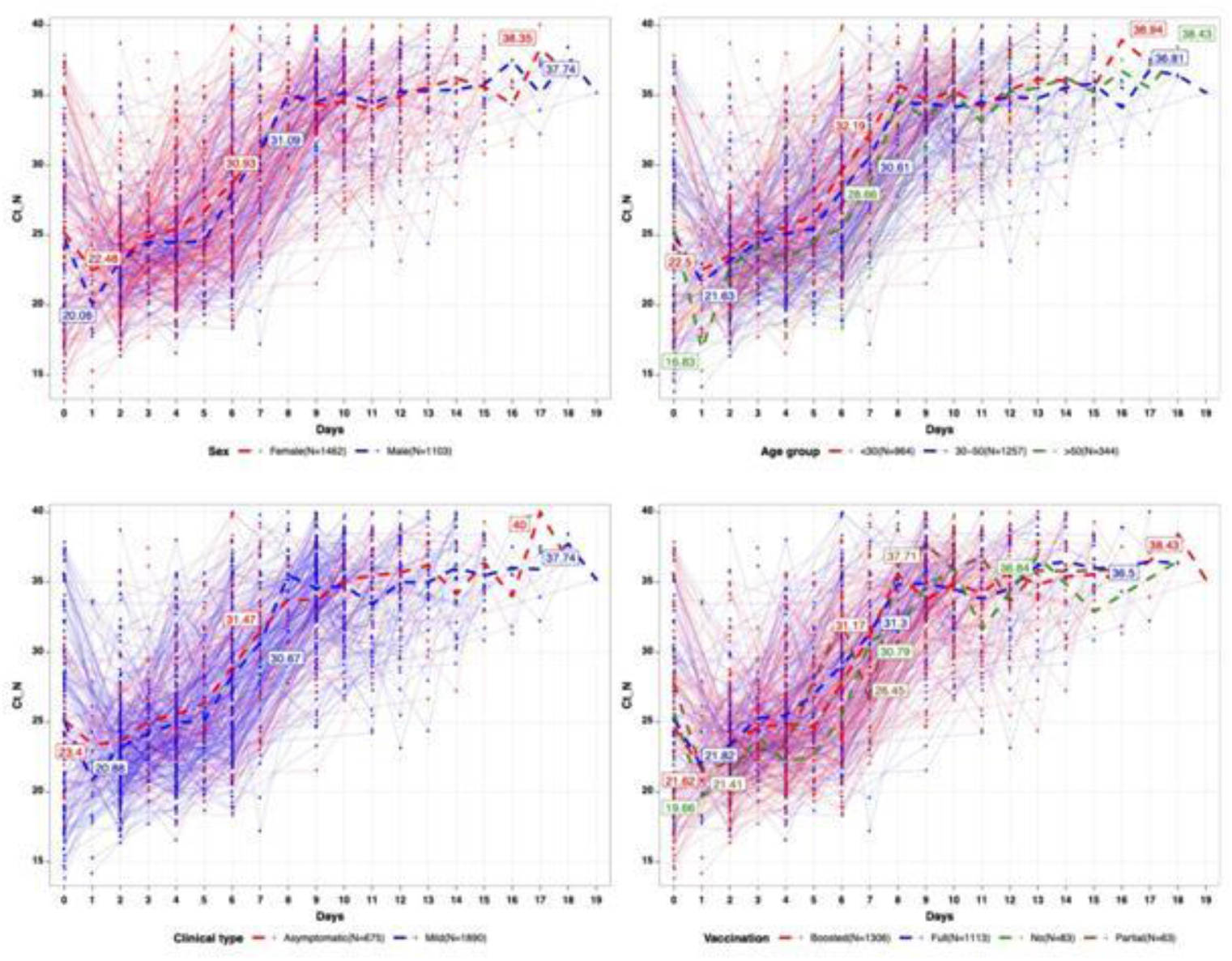
Kinetic patterns of viral shedding in patients with COVID-19 (N gene). Viral load [cycle threshold (Ct) values] detected through RT-PCR by using nasopharyngealswabs of individuals with asymptomatic or mild COVID-19 (n=374). The overall (A) and stratified by types of COVID-19 (B), sex(C), age (D), and vaccination status(E). The detection limit was Ct =40, which suggests the negative result of samples. Furthermore, patients with Ct ≥35 could be discharged from the hospital and monitored for7 days at home according to the China National guideline [8].

**Figure 4.**
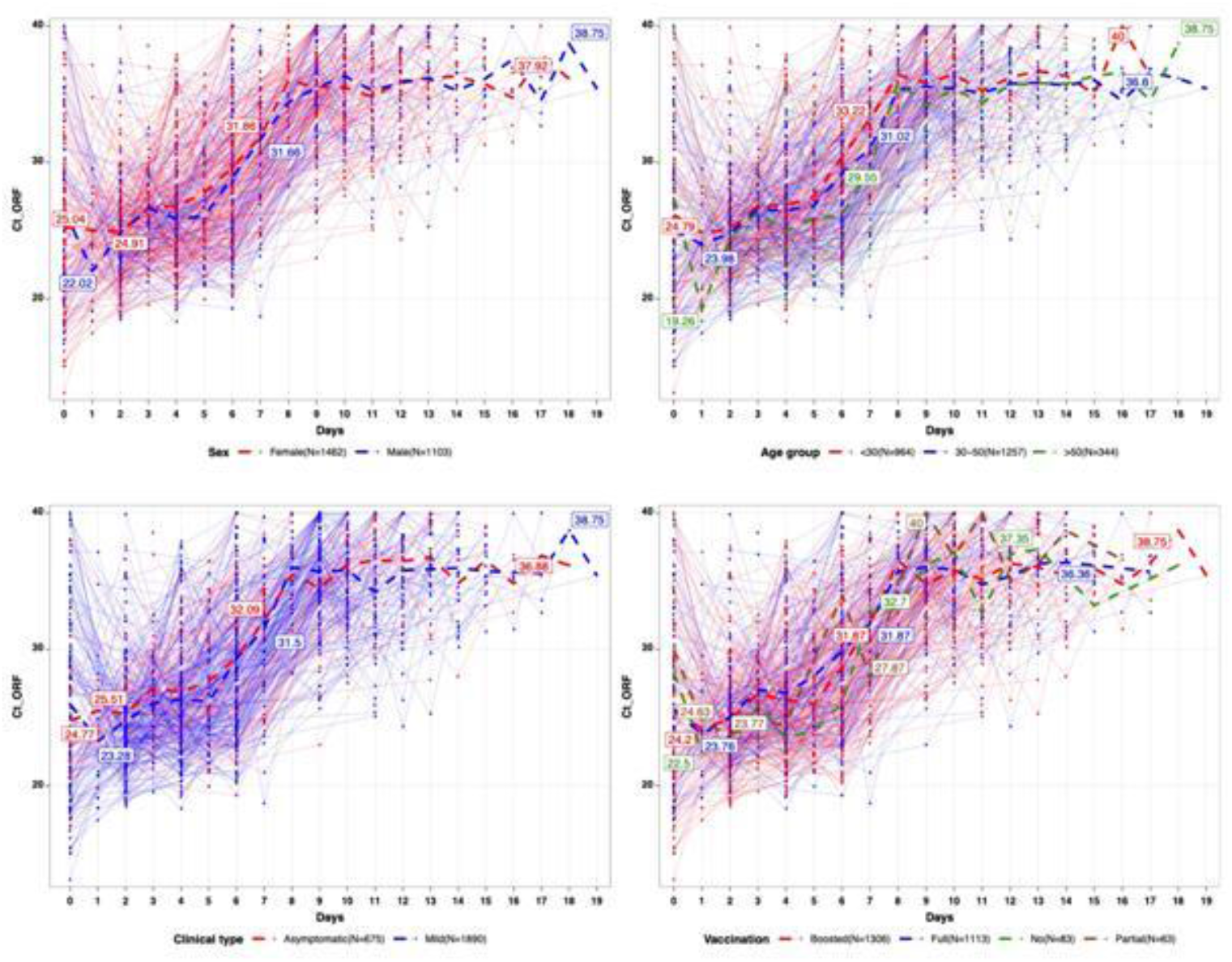
Kinetic patterns of viral shedding in patients with COVID-19 (ORF1ab gene). Viral load [cycle threshold (Ct) values] detected through RT-PCR by using nasopharyngeal swabs from individuals with asymptomatic or mild COVID-19 (n=374).

## Discussion

The highly transmissible Omicron variant spreads earlier than other variants of concern of SARS-CoV-2. Omicron has become the main variant causing infection globally [10]. China is continuing with a dynamic zero-COVID-19 policy which is based on protecting the health of the people [1].The viral shedding time directly determines the risk of COVID-19 transmission and determines the length of isolation, which plays a key role in the non-drug policy. However, the viral shedding time of the Omicron variants has not been clearly reported in China.

In 2020, our study reported the median duration of SARS-CoV-2 viral shedding to be 12.0 days in Ningbo [11], 20.0 days in survivors in Wuhan [12]or longer term in Chongqing [7].In addition, He et al. suggested that viral shedding may start 5-6 days before symptoms onset [13]. Furthermore, Wölfel et al. reported that the severity of infection might decrease significantly 8 days after symptom onset; however, they could not obtain swabs containing live virus from hospitalized COVID-19 patients [14].van Kampen et al. obtained similar results[15]. However, the Omicron variant may have shorter viral shedding durations than other variants after symptom onset or the initial positive test. According to the result of the first study from Shanghai, the viral shedding time in non-severe Omicron-infected patients infected by the Omicron variant was 6 days (IQR 3-8 days) [16].Similarly, another study on the symptomatic Omicron variant reported a median viral shedding time of 6 days in infected outpatients [17].

Recent studies have reported no difference in the viral kinetics between the infection caused by the Omicron and Delta variants [17]. The Omicron variant was still associated with continuous transmission after 8 days. SARS-CoV-2 could be detected in 50% of patients even at 5 days and in 25% patients at 8 days [17]. In addition, the Omicron variant could be detected 9 days after symptom onset in China and not at 10 days in infected patients in Japan. Furthermore, the RNA of the Omicron variant was detectable 10 days after symptom onset, while the virus could not be isolated [18]. The median viral shedding time was 10 days in 374 patients of the present study. All the patients were defined as close or sub-close contacts, and the SARS-CoV-2 nucleic acid tests were repeated, which helped in detecting the infected cases earlier than symptom onset. Therefore, we conducted an excellent real-world study by dynamically observing the viral shedding time surpass the symptom-based findings as reported [16]. On the other hand, due to the large-scale outbreak, detection of the virues by using RT-PCR might be delayed, or the admission day may not be the first day of positive SARS-CoV-2 nucleic acid tests, resulting in a shortened duration for calculating the viral shedding time.

Interestingly, the viral load in the older group (>50 years) was significantly lower than that in the younger patients, which is opposite with the results of a previous study that reported that older age was correlated with a higher viral load [19]. Furthermore, infectiousness gradually declined within 8 days of detection of the Omicron variant detection in the present study. The aforementioned evidence showed that patients infected with the Omicron variant could be managed on day 10 and the variant could no longer be transmitted. This would be criteria for discharge from shelter hospitals to be monitored at home. This would avoid placing any strain on healthcare systems during the large-scale outbreak. In addition, RT-PCR screening in the close or sub-close contacts can effectively detect the infections before symptom onset and help adopt non-drug measures, such as isolation and contact tracing.

Our study had several limitations. First, we recruited all patients with asymptomatic and mild COVID-19 from a shelter hospital but did not include moderate, severe, or critical cases. Patients with severe or critical infections might shed SARS-CoV-2 for longer periods than those with mild COVID-19 [15]. Second, the Omicron variant could not be isolated and cultured to confirm the viability of SARS-CoV-2 due to bio-security, resulting in an extended period of viral shedding. Finally, the incubation period of the Omicron variant could not be evaluated because of the large-scale outbreak.

## Conclusion

The viral shedding duration of vaccinated Omicron-infected patients with asymptomatic or mild COVID-19 (BA.2.2.1) diagnosed at the first RT-PCR was nearly 10 days. This result could provide strategies for the prevention and control of the Omicron variant-induced infection from the perspective of clinical and public health.

## Data Availability

All data produced in the present study are available upon reasonable request to the authors.

## Abbreviations

SARS-CoV-2: Severe Acute Respiratory Syndrome Coronavirus 2
COVID-19: Coronavirus disease 2019
RT-PCR: Reverse Transcription-Polymerase Chain Reaction
Ct: Cycle threshold
CDC: Center for Disease Control and Prevention
CT: Computed Tomography
VOC: Variants of Concern

## Acknowledgements

The authors are thankful to study participants.

## Author contributions

QS, and GQ conceived and designed the study. NY, DY and QS analyzed the data. YM and WS contributed data/materials. GQ and NY prepared the manuscript. All authors revised the manuscript and approved the final manuscript.

## Availability of data and materials

The dataset used and/or analyzed during the current study are available from the corresponding author on reasonable request.

## Declarations

### Ethics approval and consent to participate

This retrospective study was conducted at the Cixi Dapengshan Shelter Hospital (managed by Ningbo First Hospital, University of Ningbo), and this study was approved by the Ethics Committee of Ningbo First Hospital (approval No. 2022RS065). Written informed consent from patients was waived because this was a retrospective study.

### Consent for publication

Not applicable.

